# Peripheral immune profiles in individuals at genetic risk for amyotrophic lateral sclerosis and Alzheimer’s disease

**DOI:** 10.1101/2024.11.13.24316906

**Authors:** Laura Deecke, Olena Ohlei, David Goldeck, Jan Homann, Sarah Toepfer, Ilja Demuth, Lars Bertram, Graham Pawelec, Christina M. Lill

## Abstract

The immune system plays a crucial role in the pathogenesis of neurodegenerative diseases. Here, we explored whether blood immune cell profiles are already altered in healthy individuals with a genetic predisposition to amyotrophic lateral sclerosis (ALS) or Alzheimer’s disease (AD).

Using multicolor flow cytometry, we analyzed 92 immune cell phenotypes in the blood of 448 healthy participants from the Berlin Aging Study II. We calculated polygenic risk scores (PGS) using genome-wide significant SNPs from recent large genome-wide association studies on ALS and AD. Linear regression analyses were then performed of the immune cell types on the PGS in both the overall sample and a subgroup of older participants (>60 years).

While we did not find any significant associations between immune cell subtypes and ALS and AD PGS when controlling for the false discovery rate (FDR=0.05), we observed several nominally significant results (p<0.05) with consistent effect directions across strata. The strongest association was observed with CD57+ CD8+ early-memory T cells and ALS risk (p=0.006). Other immune cell subtypes associated with ALS risk included PD-1+ CD8+ and CD57+ CD4+ early-memory T cells, non-classical monocytes, and myeloid dendritic cells. For AD, naïve CD57+ CD8+ T cells, and mature NKG2A+ natural killer cells showed nominally significant associations.

We did not observe immune cell changes in individuals with high genetic risk for ALS or AD, suggesting immune alterations may arise later in disease progression. Additional studies are required to validate our nominally significant findings.

## Introduction

The development of neurodegenerative diseases (ND) arises from a complex interplay between genetic and environmental factors. Amyotrophic lateral sclerosis (ALS) is characterized by the degeneration of both first and second motor neurons resulting in severe symptoms of muscle weakness and ultimately neuromuscular respiratory failure, making it one of the most fatal forms of ND^1^. Alzheimer’s Disease (AD), the most common ND, is characterized by the accumulation of amyloid-beta and tau proteins in the brain, ultimately leading to cognitive decline and dementia^2^. In this context, increasing evidence points to a critical role of the immune system in the pathogenesis of NDs^3^. For instance, alterations of different T cell subsets in the central nervous system as well as in the periphery have been reported in prevalent ALS^4^ and AD^5^ patients compared to healthy controls. Recently, our group investigated possible alterations of CD8+ TEMRA cells in healthy individuals with high genetic risk for AD^6^ based on prior reports suggesting differential levels of these cells in pre-clinical AD cases^7^. Although these analyses did not confirm a role for CD8+ TEMRA cells in the early pre-clinical phase of AD, the fact that changes in immune cell composition associated with AD or ALS are likely to precede the clinical onset of these diseases by years, prompted us to investigate early immune cell alterations using a more comprehensive approach. To this end, we investigated whether healthy individuals at high genetic risk for these diseases show alterations in relevant immune cell subtypes. Specifically, we calculated polygenic risk scores (PGS) for ALS and AD and examined their associations with 92 immune cell subtypes in ~450 individuals from the Berlin Aging Study II (BASE-II).

## Methods

### Study participants

A total of 448 European individuals recruited as part of BASE-II were included in this study (described in ref. ^6^). BASE-II is a multi-institutional longitudinal cohort study from the larger Berlin area in Germany that investigates determinants and mechanisms of aging (described in detail elsewhere^8^). The effective dataset investigated here consists of individuals from two age groups, comprising 309 older adults (60% females, median age: 69 years, age range:60-82 years) and 139 younger adults (59% females, median age: 29, age range: 23-35, **Supplementary Figure 1**). Individuals with neurodegenerative diseases were excluded. All individuals were immunologically healthy (no fever or immune system-related diseases), had no diagnosis of neurodegenerative diseases, and were cognitively unimpaired (previously described ref. ^7^), and quality-controlled (QC) genetic as well as immune cell data were available.

### Generation and processing of immune cell and genome-wide SNP data

Peripheral blood mononuclear cells were isolated and analyzed by multi-colour flow cytometry as described previously^6,9^. Briefly, two different immune cell antibody panels were used to distinguish between various immune cell subtypes, i.e. subsets of T cells, B cells, natural killer cells or monocytes (**Supplementary Table 1-2**). A 3 laser BD LSRII (BD biosciences) flow cytometer and DIVA6 software were utilized for the acquisition of the data, analyzed thereafter using FlowJo version 7.5 as described previously^6,9^. QC’ed immune cells were quantified as proportions of the major immune cell types and distributions were transformed where appropriate (based on visual inspections) using one of the following approaches: log10, root, log(100-x) or square^10^ (**Supplementary Table 1**). As a final step, the data were z-transformed.

The genome-wide SNP data were generated and processed as described before^6^. In short, the Affymetrix Array 6.0 was used for genotyping, followed by imputation of unmeasured genotypes using the Haplotype Reference Consortium reference panel and finalized using standard QC procdures^6^. After filtering for a minor allele frequency (MAF) threshold of <0.01 and an imputation quality threshold of <0.3, we obtained a final set of 7,512,709 SNPs. As the imputation of the established AD *APOE* risk SNP e4 (rs429358) was suboptimal in this dataset (imputation r2=0.64), this SNP was directly genotyped using “Taqman” technology (Thermo Fisher Scientific, Inc.)^6^.

The calculation of the PGS for ALS was based on 11 index SNPs that showed genome-wide statistical significance in European individuals from a recent large genome-wide association study (GWAS) on ALS^11^ (**Supplementary Table 3, Supplementary Figure 2**). By including these 11 SNPs, we relaxed the minor allele frequency threshold for one SNP (rs80265967, MAF=0.00039) to maximize the number of SNPs included in the PGS. As sensitivity analysis we re-calculated an additional PGS and subsequent association statistics excluding this SNP (**Supplementary Table 3**). PGS calculations for AD were based on 70 genome-wide significant SNPs (α=5.0E-8) from a recent GWAS on AD^12^. Of note, the beta estimate and p value of the *APOE* e4 SNP were taken from an earlier AD GWAS^13^ by the same group, because it was excluded from the summary statistics in ref. ^12^ (**Supplementary Table 4**). Due to missing data of the *APOE* e4 genotype in some BASE-II participants, some individuals had to be excluded from the analyses (n=18), resulting in a maximum of 430 individuals included in the AD analyses (**Supplementary Figure 2**). The PGS was z-transformed prior to statistical analyses.

### Statistical analyses

Linear regression analyses using the z-transformed PGS as exposure and the z-transformed immune cell proportions as outcome were performed, while adjusting for sex, the age group, and the first four genetic principal components to account for potential population substructures. The analyses were conducted using the ‘lm’ function in R. The main analyses were performed on the full sample and the subgroup of older BASE-II participants. Additionally, exploratory analyses stratifying for sex were performed on all nominally significant results (α=0.05). Sensitivity analyses included an adjustment for cytomegalovirus (CMV) status and utilization of a modified PGS for ALS. The threshold for the false discovery rate (FDR) was set to 5%.

## Results

The linear regression analyses of PGS on 92 immune cell subtypes in the full sample and the subgroup of older individuals did not yield significant signals at an FDR of 5%, neither for ALS nor for AD (**Supplementary Figure 3)**. Despite the lack of study-wide significant findings, we observed several nominally significant (α=0.05) – and hence potentially relevant – signals:

For ALS, five cell types passed the nominal threshold in analyses of the full BASE-II sample (**Figure 1**): This included the T cell subsets CD57+ CD8+ early-memory T cells (p=0.006), PD-1+ CD8+ T cells (p=0.024), and CD57+ CD4+ early-memory T cells (p=0.038), as well as non-classical monocytes (p=0.015) and myeloid dendritic cells (p=0.047) with variances explained (ΔR2) ranging from 2.1% to 1.1% (**Table 1**). Notably, CD57+ CD8+ early-memory T cells exhibited consistent effect estimates across all analyses, and importantly, showed the strongest effect estimates in men with the PGS explaining 7% of the variance in immune cell distribution in this subset (p=0.001). The same cell type also reached nominal significance when analyses were restricted to the older age group (ΔR2=2.4%, p=0.012). In addition, PD-1+ CD8+ T cells, also exhibiting consistent effect estimates across strata, showed nominally significant association not only in the full sample but also in the subset of men (ΔR2=2.6%, p=0.041; **Table 1**).

**Figure 1.**
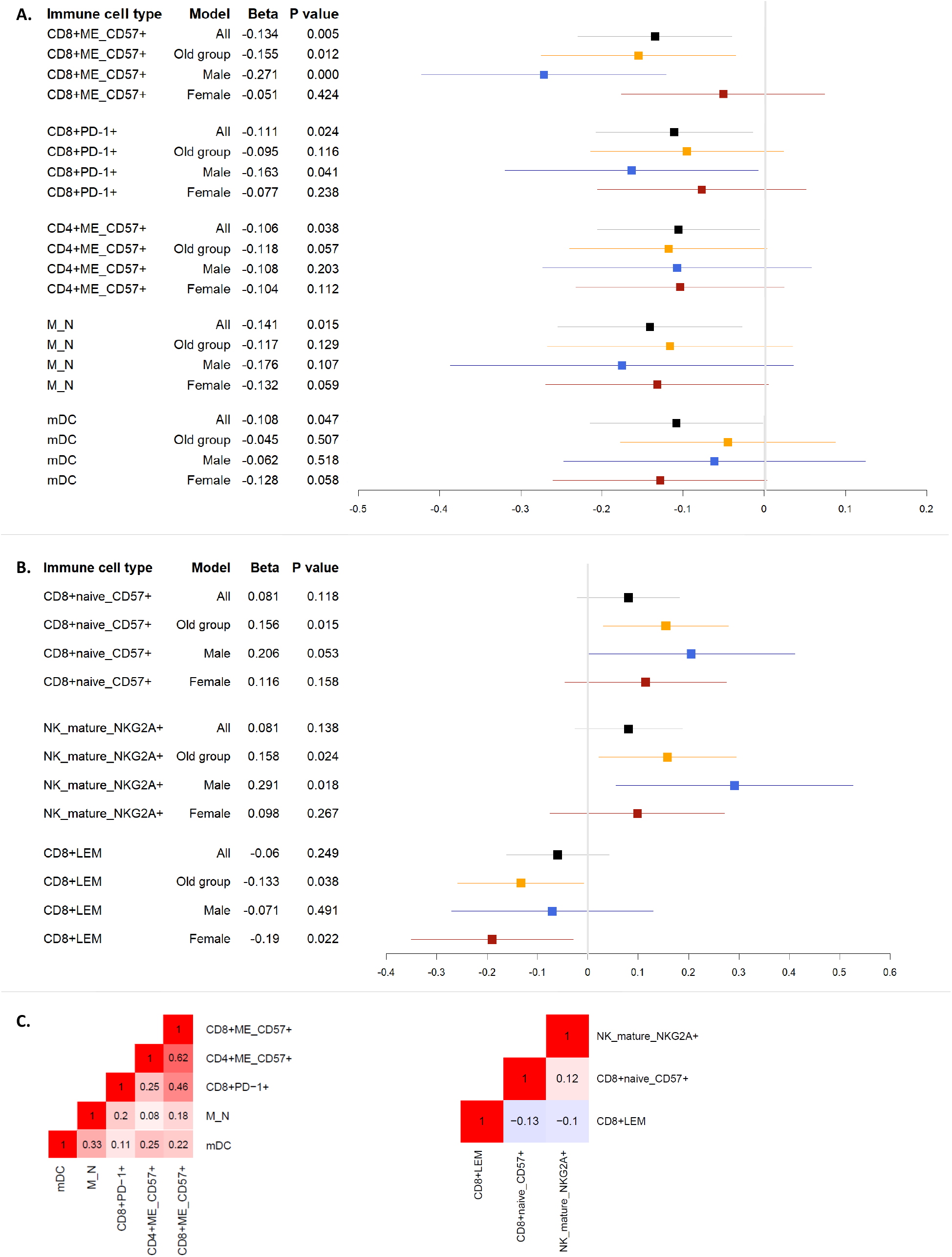
Nominally significant linear regression results of neurodegenerative disease polygenic risk score with immune cell distributions in the blood of healthy BASE-II participants. Legend. This forest plot displays beta values and 95% confidence intervals of nominal significant association results linking amyotrophic laterals sclerosis (A) and Alzheimer’s disease (B) and levels immune cell subtypes in blood as well as correlations between the corresponding nominally significant cell types per disease (C).

**Table 1.** Immune cells showing nominally significant associations with genetic risk profiles of amyotrophic lateral sclerosis and Alzheimer’s disease.

| | Immune cell type | Model | Beta | $\Delta R^2$ | P |
| --- | --- | --- | --- | --- | --- |
| **ALS** | **CD57+ CD8+ early-memory T cells** | **All** | -0.135 | 0.018 | **0.006** |
|  | **CD57+ CD8+ early-memory T cells** | **Old group** | -0.155 | 0.024 | **0.012** |
|  | CD57+ CD8+ early-memory T cells | Female | -0.051 | 0.003 | 0.424 |
|  | **CD57+ CD8+ early-memory T cells** | **Male** | -0.271 | 0.072 | **0.001** |
|  | **PD-1+ CD8+ T cells** | **All** | -0.111 | 0.012 | **0.024** |
|  | PD-1+ CD8+ T cells | Old group | -0.095 | 0.009 | 0.116 |
|  | PD-1+ CD8+ T cells | Female | -0.077 | 0.006 | 0.238 |
|  | **PD-1+ CD8+ T cells** | **Male** | -0.163 | 0.026 | **0.041** |
|  | **Non-classical monocytes** | **All** | -0.141 | 0.021 | **0.015** |
|  | Non-classical monocytes | Old group | -0.117 | 0.015 | 0.129 |
|  | Non-classical monocytes | Female | -0.133 | 0.018 | 0.059 |
|  | Non-classical monocytes | Male | -0.176 | 0.031 | 0.107 |
|  | **CD57+ CD4+ early-memory T cells** | **All** | -0.106 | 0.011 | **0.038** |
|  | CD57+ CD4+ early-memory T cells | Old group | -0.118 | 0.014 | 0.057 |
|  | CD57+ CD4+ early-memory T cells | Female | -0.104 | 0.011 | 0.112 |
|  | CD57+ CD4+ early-memory T cells | Male | -0.108 | 0.011 | 0.203 |
|  | **Myeloid dendritic cells** | **All** | -0.108 | 0.012 | **0.047** |
|  | Myeloid dendritic cells | Old group | -0.045 | 0.002 | 0.507 |
|  | Myeloid dendritic cells | Female | -0.128 | 0.017 | 0.058 |
|  | Myeloid dendritic cells | Male | -0.062 | 0.004 | 0.518 |
| **AD** | Naïve CD57+ CD8+ T cells | All | 0.081 | 0.007 | 0.118 |
|  | **Naïve CD57+ CD8+ T cells** | **Old group** | 0.156 | 0.024 | **0.015** |
|  | Naïve CD57+ CD8+ T cells | Female | 0.116 | 0.013 | 0.158 |
|  | Naïve CD57+ CD8+ T cells | Male | 0.206 | 0.039 | 0.053 |
|  | Mature NKG2A+ natural killer cells | All | 0.081 | 0.007 | 0.138 |
|  | **Mature NKG2A+ natural killer cells** | **Old group** | 0.158 | 0.024 | **0.024** |
|  | Mature NKG2A+ natural killer cells | Female | 0.098 | 0.009 | 0.267 |
|  | **Mature NKG2A+ natural killer cells** | **Male** | 0.291 | 0.079 | **0.018** |
|  | CD8+ late effector memory T cells | All | -0.060 | 0.004 | 0.249 |
|  | **CD8+ late effector memory T cells** | **Old group** | -0.133 | 0.017 | **0.038** |
|  | **CD8+ late effector memory T cells** | **Female** | -0.112 | 0.013 | **0.022** |
|  | CD8+ late effector memory T cells | Male | -0.071 | 0.005 | 0.491 |

**Legend.** This table visualizes the results on all immune cell analyses that show nominally significant ( $\alpha=0.05$ ) association in a least one analysis (i.e., analyzing all or restricting to the older group). The linear regression analyses were performed across 448 healthy individuals from the Berlin Aging Study II (BASE-II) using polygenic scores (PGS) for amyotrophic lateral sclerosis (ALS) and Alzheimer's disease (AD) as exposure and immune cell proportions as outcome. Beta = effect estimate of regression analyses of z-transformed immune cell distributions on z-transformed polygenic risk scores. $\Delta R^2$ = variance of immune cell distribution explained by the polygenic risk score.

For AD, we did not observe any trends of association (α=0.05) in the full BASE-II dataset. However, upon restricting our investigation to the older subsample, i.e., that closer to a potential clinical onset of AD, showed trends for an association of higher genetic AD risk with subsets of CD8+ T cells and natural killer cells (**Figure 1**). Specifically, a higher PGS was associated with increased naïve CD57+ CD8+ T cells (p=0.015), increased mature NKG2A+ natural killer cells (p=0.024), and decreased CD8+ late effector memory T cells (p=0.038) with ΔR2 ranging from 2.4% to 1.7%. The latter two associations are driven by the male and female subgroup, respectively (**Table 1**).

Lastly, none of the results changed appreciably in the sensitivity analyses adjusting for CMV or using the alternative PGS for ALS (**Supplementary Table 5-6**).

## Discussion

In this study, we performed a comprehensive investigation as to whether alterations in peripheral blood immune cell composition are evident in healthy participants at high genetic risk for AD or ALS. While none of the associations reached study-wide significance in ~450 BASE-II participants after FDR control, we observed several nominally significant associations between genetic ALS and AD risk and various immune cell subsets. These include subsets of CD4+, CD8+ T cells, monocytes, and myeloid dendritic cells for ALS as well as subsets of CD8+ T cells and natural killer cells for AD. To our knowledge, this is the first study to analyze such a broad spectrum of immune cell subtypes in healthy individuals at high risk of ALS or AD.

Notably, overall the nominally significant results showed consistent associations across analyzed strata. The most compelling results for ALS suggest a decrease in CD57+ CD8+ early-memory T cells and in PD-1+ CD8+ T cells. Interestingly, a recent landmark study investigated both effector and memory CD8+ T cells in human monogenic ALS patients with *SETX* mutations as well as PD-1+ CD8+ T cells in mice^14^. The authors found a general decrease of CD8+ effector and memory T cells, which is in agreement with our observations. While they did not show data for PD-1+ CD8+ T cells in humans, they described differential proportions of these cells in a murine ALS model. However, effect directions differed in mice vs our human data, potentially reflecting the limited comparability between mouse models and humans^14^.

In individuals at high genetic risk for AD, the most noteworthy immune cell alterations highlighted by our analyses relate to increased naïve CD57+ CD8+ T cells and mature NKG2A+ natural killer cells. Consistent effect directions were observed for the main and all subgroup analyses. Notably the vast majority of studies investigating immune cells analyzed prevalent ALS and AD cases. While we are not aware of specific investigations of naïve CD57+ CD8+ T cells in AD, a number of studies reported on CD8+ T cell abnormalities in AD patients^15^. A role for natural killer cells in the pathogenesis of AD has been reported repeatedly^15,16^, however their exact mechanisms including mature NKG2A+ natural killer cells remain elusive at this point in time. It is noteworthy that we found differences of immune cells for individuals of AD risk only in the smaller older age but not in the overall group, a pattern which we did not observe for ALS risk. This difference may be attributable to the approximately ten year later age of onset of AD compared to ALS.

The primary strengths of this study are an extensive and thorough clinical examination of all BASE-II participants, in-depth immune cell phenotyping, and a comparatively large sample size. Potential limitations of our work include limited statistical power to detect minor effects, the missing consideration of lifestyle and environmental ND risk factors, and the lack of data in non-European individuals, as discussed in previous work from our group^17^. In that study, we examined alterations in immune cell types in healthy individuals with a high genetic risk for Parkinson’s disease^17^. Together with that study, our findings suggest that changes in immune cell composition may not manifest in healthy individuals with an increased genetic risk of the investigated NDs. Given the well-established role of the immune system response in ND pathogenesis^3^, this suggests that immune cell alterations become manifest at later stages of disease development. Notwithstanding, our work highlights interesting trends of alterations in several innate and adaptive immune cell subtypes, which warrant further investigation in future research.

## Author Contributions

Conceptualization: C.M.L.; methodology: L.D., C.M.L.; formal analysis or interpretation of the data: L.D., O.O., J.H., C.M.L.; data acquisition: D.G., I.D., G.P., L.B.; writing—original draft preparation: L.D., C.M.L.; writing—review and editing: D.G., O.O., J.H., I.D., L.B., G.P.; supervision, C.M.L. All authors have read and agreed to the final version of the manuscript.

## Data Availability

All summary statistics have been made available in the supplementary material of this manuscript. Raw and source data are available upon reasonable request. Interested researchers may contact the scientific BASE-II coordinator, Ludmila Müller. Additional information is available on the BASE-II website: https://www.base2.mpg.de/7549/data-documentation.

## Acknowledgements

We are grateful to all BASE-II participants.

## Ethical approval

All BASE-II participants provided written informed consent before participation and the study was conducted in accordance with the Declaration of Helsinki and approved by the Ethics Committee of the Charité-Universitätsmedizin Berlin – approval number EA2/029/09.

