## Supplementary material for "Peripheral immune profiles in individuals at genetic risk for amyotrophic lateral sclerosis and Alzheimer’s disease": Deecke_immuneAD_ALS_PGS_supplementary_material_final-Cells.pdf

**Supplementary Figure 1.** Age distribution in the Berlin Aging Study II

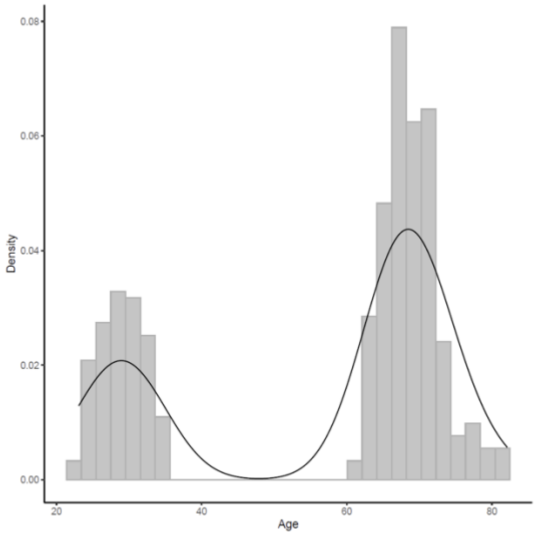


**Supplementary Figure 2.** Distributions of the polygenic risk scores for amyotrophic lateral sclerosis (A) and Alzheimer's disease (B)

2A.

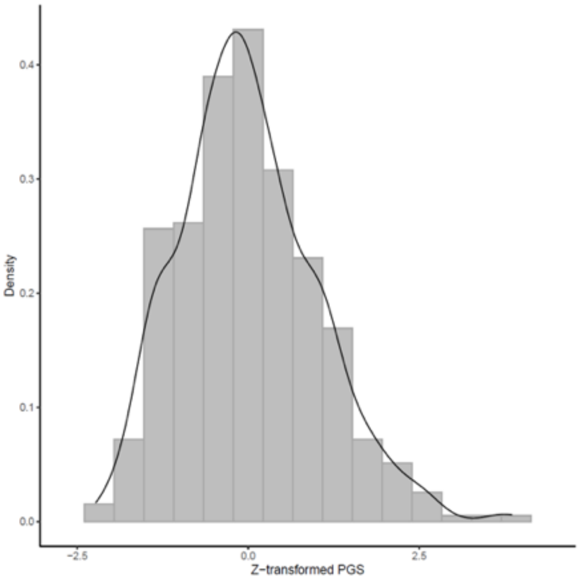


2B.

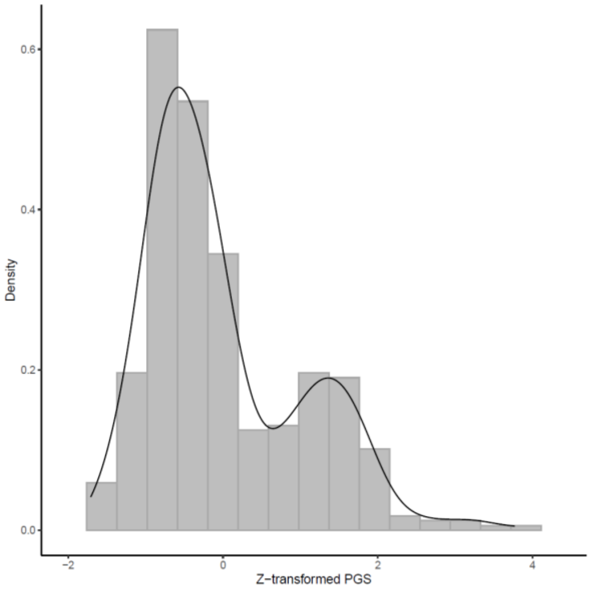

**Supplementary Figure 3.** Effect size estimates of the polygenic risk scores for amyotrophic lateral sclerosis (ALS) and for Alzheimer's disease (AD) on all immune cell distributions analyzed

#### 3A. ALS, all participants

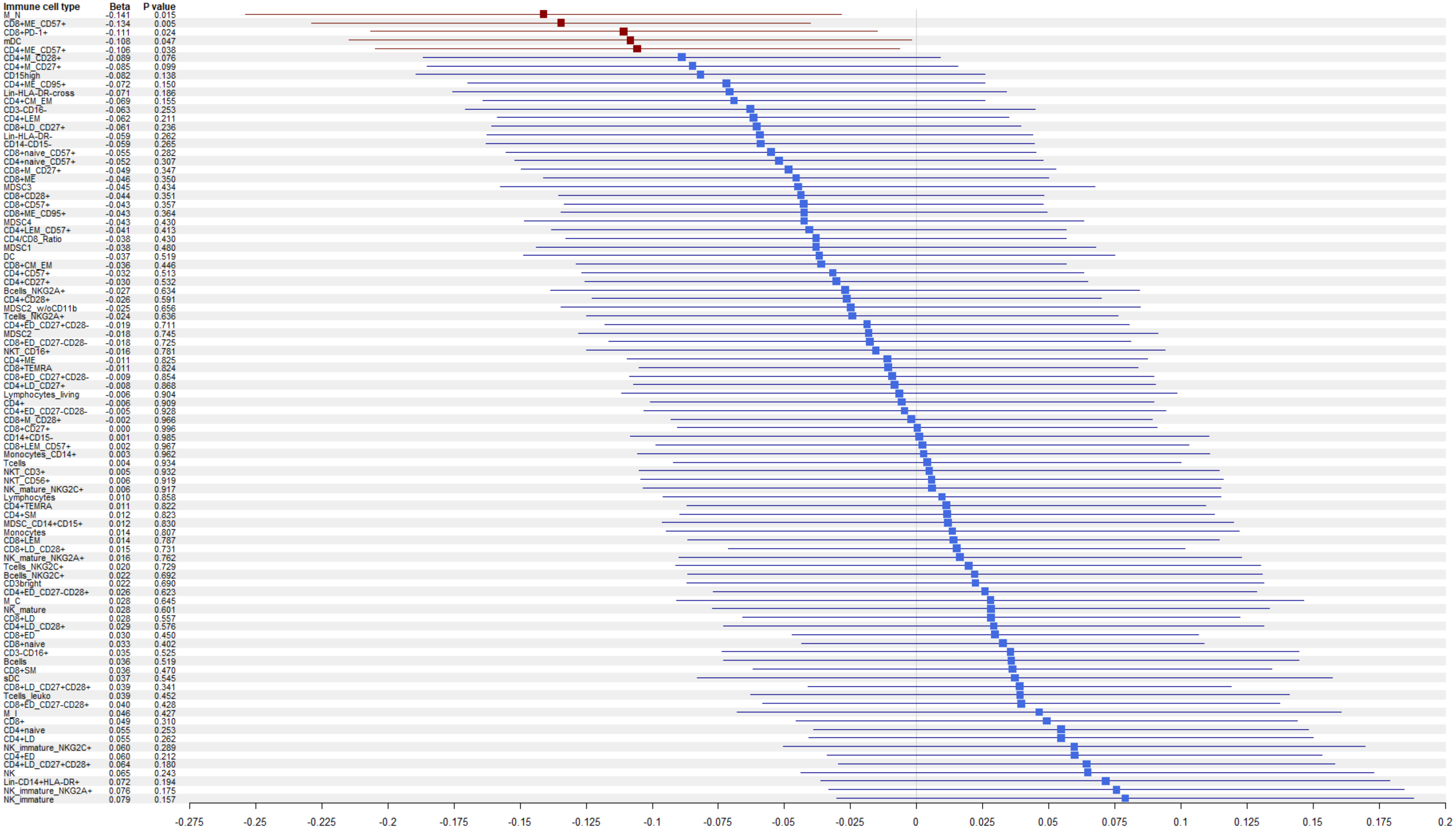

### 3B. ALS, older age group

| Immune cell type | Beta | P value |
|---|---|---|
| CD8+ME_CD57+ | -0.155 | 0.012 |
| CD4+ME_CD57+ | -0.118 | 0.057 |
| M_N | -0.117 | 0.129 |
| CD4+M_CD27+ | -0.097 | 0.118 |
| CD8+PD-1+ | -0.095 | 0.116 |
| CD4+M_CD28+ | -0.087 | 0.160 |
| NKT_CD16+ | -0.085 | 0.213 |
| CD8+naive_CD57+ | -0.082 | 0.188 |
| CD4+naive_CD57+ | -0.074 | 0.236 |
| CD8+LD_CD27+ | -0.071 | 0.258 |
| CD15high | -0.069 | 0.313 |
| CD8+M_CD27+ | -0.062 | 0.320 |
| DC | -0.055 | 0.452 |
| CD8+CD28+ | -0.052 | 0.389 |
| CD8+CD57+ | -0.052 | 0.381 |
| CD4+LEM | -0.048 | 0.433 |
| CD4/CD8_Ratio | -0.047 | 0.446 |
| MDSC2_w/oCD11b | -0.047 | 0.492 |
| CD8+ME | -0.046 | 0.449 |
| CD3-CD16- | -0.046 | 0.502 |
| MDSC3 | -0.045 | 0.525 |
| mDC | -0.045 | 0.507 |
| CD8+ME_CD95+ | -0.044 | 0.472 |
| CD4+CM_EM | -0.043 | 0.483 |
| CD8+TEMRA | -0.042 | 0.502 |
| CD14-CD15- | -0.041 | 0.547 |
| MDSC2 | -0.041 | 0.548 |
| CD14+CD15- | -0.041 | 0.553 |
| CD4+ME_CD95+ | -0.039 | 0.536 |
| CD4+SM | -0.039 | 0.545 |
| CD4+LEM_CD57+ | -0.039 | 0.533 |
| CD8+CM_EM | -0.034 | 0.578 |
| CD4+ED_CD27+CD28- | -0.031 | 0.616 |
| CD8+ED_CD27-CD28- | -0.029 | 0.633 |
| Lin-HLA-DR-cross | -0.026 | 0.702 |
| CD8+LD_CD28+ | -0.024 | 0.693 |
| Lin-HLA-DR- | -0.020 | 0.771 |
| CD4+ED_CD27-CD28+ | -0.019 | 0.766 |
| CD8+CD27+ | -0.018 | 0.766 |
| CD4+ | -0.018 | 0.776 |
| MDSC4 | -0.012 | 0.856 |
| CD4+CD57+ | -0.012 | 0.850 |
| CD8+M_CD28+ | -0.012 | 0.851 |
| Bcells_NKG2A+ | -0.010 | 0.887 |
| NK_mature_NKG2A+ | -0.010 | 0.887 |
| CD4+LD_CD27+ | -0.009 | 0.882 |
| CD4+CD27+ | -0.007 | 0.906 |
| CD4+LD_CD28+ | -0.006 | 0.923 |
| CD8+ED_CD27+CD28- | -0.005 | 0.934 |
| CD4+CD28+ | -0.004 | 0.942 |
| CD3-CD16+ | -0.004 | 0.958 |
| CD8+SM | 0.000 | 0.994 |
| Monocytes | 0.001 | 0.989 |
| CD4+ED_CD27-CD28- | 0.003 | 0.961 |
| Tcells_NKG2A+ | 0.005 | 0.941 |
| Monocytes_CD14+ | 0.007 | 0.922 |
| MDSC1 | 0.007 | 0.918 |
| CD8+ED_CD27-CD28+ | 0.009 | 0.886 |
| CD8+LEM_CD57+ | 0.011 | 0.859 |
| NK_mature_NKG2C+ | 0.017 | 0.798 |
| CD8+LEM | 0.023 | 0.720 |
| CD8+LD | 0.024 | 0.708 |
| NK_mature | 0.025 | 0.715 |
| CD4+naive | 0.025 | 0.678 |
| Lymphocytes_living | 0.026 | 0.706 |
| Tcells | 0.026 | 0.680 |
| NKT_CD3+ | 0.028 | 0.687 |
| NKT_CD56+ | 0.030 | 0.667 |
| NK | 0.031 | 0.663 |
| CD4+TEMRA | 0.031 | 0.613 |
| MDSC_CD14+CD15+ | 0.032 | 0.635 |
| CD4+ED | 0.033 | 0.588 |
| CD4+ME | 0.035 | 0.586 |
| CD4+LD_CD27+CD28+ | 0.035 | 0.569 |
| CD3bright | 0.037 | 0.585 |
| CD4+LD | 0.038 | 0.532 |
| M_I | 0.040 | 0.595 |
| CD8+ED | 0.040 | 0.496 |
| CD8+naive | 0.041 | 0.490 |
| Tcells_NKG2C+ | 0.041 | 0.545 |
| Lymphocytes | 0.048 | 0.479 |
| CD8+LD_CD27+CD28+ | 0.050 | 0.414 |
| M_C | 0.052 | 0.482 |
| CD8+ | 0.053 | 0.393 |
| sDC | 0.053 | 0.477 |
| NK_immature | 0.067 | 0.331 |
| NK_immature_NKG2A+ | 0.067 | 0.327 |
| Tcells_leuko | 0.070 | 0.302 |
| Bcells_NKG2C+ | 0.071 | 0.298 |
| Lin-CD14+HLA-DR+ | 0.072 | 0.294 |
| Bcells | 0.076 | 0.252 |
| NK_immature_NKG2C+ | 0.115 | 0.096 |

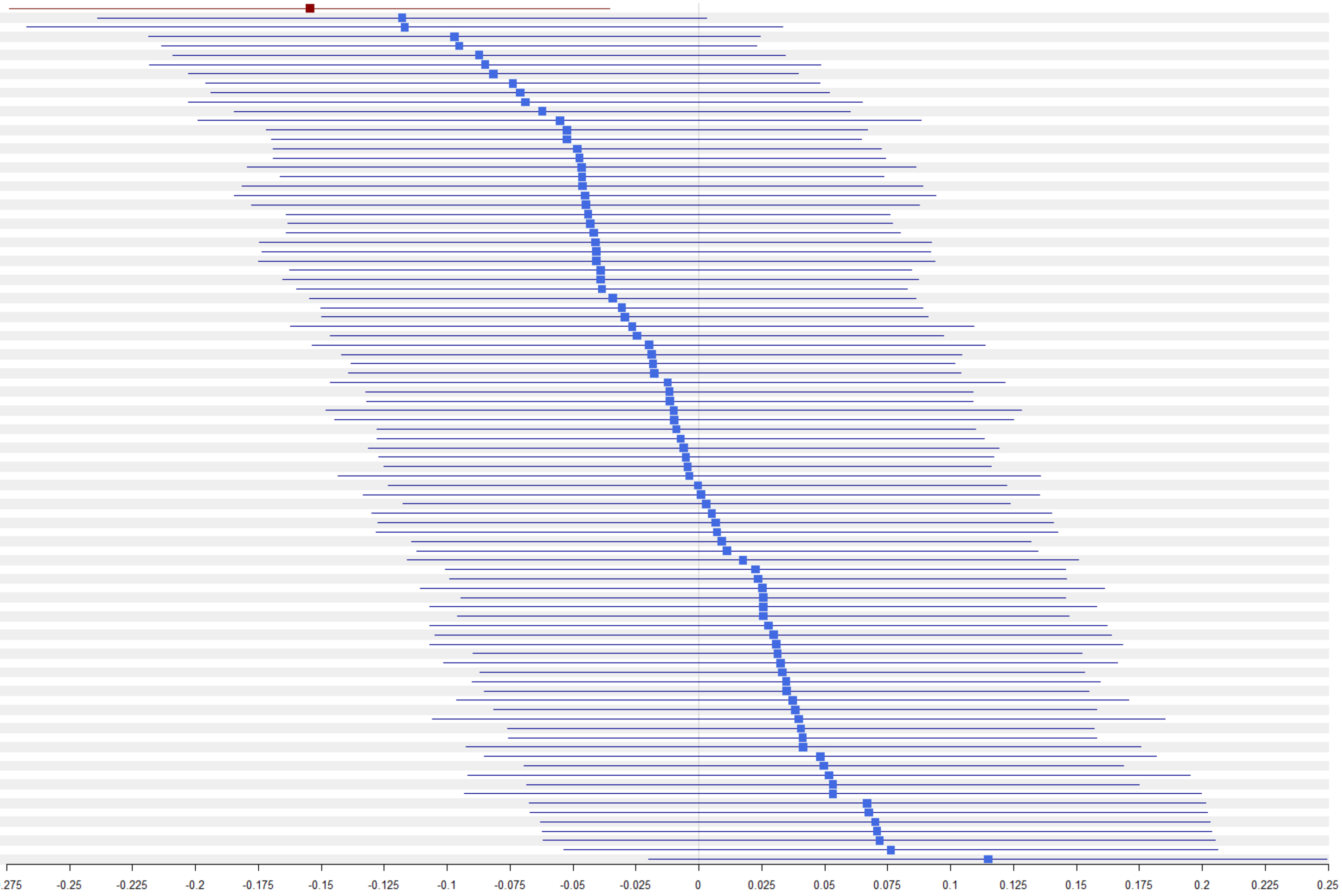

### 3C. AD, all participants

| Immune cell type | Beta | P value |
|---|---|---|
| Tcells_NKG2C+ | -0.102 | 0.071 |
| NKT_CD56+ | -0.078 | 0.172 |
| NKT_CD3+ | -0.065 | 0.252 |
| CD8+LEM_CD57+ | -0.064 | 0.217 |
| CD8+LEM | -0.060 | 0.249 |
| CD4+CD28+ | -0.053 | 0.296 |
| CD4+ED_CD27+CD28- | -0.051 | 0.322 |
| CD4+SM | -0.051 | 0.329 |
| Bcells_NKG2C+ | -0.051 | 0.361 |
| CD4+LD_CD27+ | -0.046 | 0.380 |
| CD8+ED_CD27-CD28- | -0.042 | 0.418 |
| CD8+CD28+ | -0.042 | 0.385 |
| CD8+CM_EM | -0.040 | 0.407 |
| CD4+ED_CD27-CD28+ | -0.036 | 0.496 |
| CD8+CD27+ | -0.032 | 0.505 |
| CD4+M_CD27+ | -0.029 | 0.577 |
| Tcells | -0.028 | 0.572 |
| CD4+TEMRA | -0.028 | 0.586 |
| CD4+ED_CD27-CD28- | -0.026 | 0.608 |
| CD4/CD8_Ratio | -0.026 | 0.604 |
| CD8+TEMRA | -0.025 | 0.606 |
| CD4+ME | -0.025 | 0.622 |
| Monocytes | -0.020 | 0.711 |
| CD8+ED_CD27-CD28+ | -0.020 | 0.695 |
| CD3bright | -0.018 | 0.747 |
| Lymphocytes_living | -0.017 | 0.756 |
| CD4+LD_CD27+CD28+ | -0.014 | 0.769 |
| CD14+CD15- | -0.012 | 0.826 |
| Lymphocytes | -0.012 | 0.820 |
| NK_mature_NKG2C+ | -0.010 | 0.859 |
| CD8+CD57+ | -0.009 | 0.854 |
| sDC | -0.008 | 0.898 |
| M_N | -0.008 | 0.895 |
| CD4+ED | -0.006 | 0.904 |
| CD4+ | -0.005 | 0.925 |
| CD4+CD57+ | -0.004 | 0.928 |
| CD4+LEM | -0.004 | 0.938 |
| CD4+LD | -0.003 | 0.955 |
| NK | -0.002 | 0.968 |
| CD4+naive | -0.002 | 0.969 |
| CD8+ED_CD27+CD28- | -0.001 | 0.981 |
| CD4+LEM_CD57+ | -0.001 | 0.982 |
| mDC | 0.000 | 0.993 |
| CD8+PD-1+ | 0.001 | 0.980 |
| CD8+ME | 0.001 | 0.979 |
| Tcells_leuko | 0.003 | 0.959 |
| CD8+LD_CD28+ | 0.003 | 0.953 |
| M_C | 0.003 | 0.965 |
| CD4+LD_CD28+ | 0.004 | 0.943 |
| Bcells | 0.004 | 0.947 |
| CD3-CD16+ | 0.006 | 0.916 |
| MDSC2 | 0.006 | 0.915 |
| CD8+ME_CD95+ | 0.007 | 0.879 |
| MDSC2_w/oCD11b | 0.010 | 0.864 |
| CD4+CD27+ | 0.012 | 0.804 |
| M_I | 0.015 | 0.803 |
| CD8+LD | 0.015 | 0.761 |
| NK_mature | 0.018 | 0.745 |
| Monocytes_CD14+ | 0.018 | 0.743 |
| Lin-CD14+HLA-DR+ | 0.020 | 0.723 |
| CD4+naive_CD57+ | 0.020 | 0.698 |
| CD8+ED | 0.021 | 0.600 |
| CD8+M_CD28+ | 0.023 | 0.632 |
| CD4+CM_EM | 0.023 | 0.642 |
| CD8+LD_CD27+CD28+ | 0.028 | 0.508 |
| CD8+naive | 0.028 | 0.480 |
| CD8+ | 0.031 | 0.536 |
| MDSC_CD14+CD15+ | 0.031 | 0.574 |
| CD8+LD_CD27+ | 0.034 | 0.513 |
| CD4+ME_CD95+ | 0.034 | 0.498 |
| CD8+M_CD27+ | 0.036 | 0.502 |
| DC | 0.039 | 0.502 |
| MDSC4 | 0.039 | 0.470 |
| CD8+ME_CD57+ | 0.040 | 0.424 |
| CD3-CD16- | 0.041 | 0.466 |
| Lin-HLA-DR- | 0.042 | 0.432 |
| CD4+ME_CD57+ | 0.043 | 0.406 |
| CD8+SM | 0.043 | 0.399 |
| Tcells_NKG2A+ | 0.046 | 0.373 |
| NK_immature_NKG2C+ | 0.047 | 0.409 |
| CD15high | 0.052 | 0.355 |
| CD4+M_CD28+ | 0.053 | 0.299 |
| Lin-HLA-DR-cross | 0.054 | 0.326 |
| CD14-CD15- | 0.055 | 0.302 |
| MDSC1 | 0.067 | 0.219 |
| NK_immature | 0.071 | 0.207 |
| Bcells_NKG2A+ | 0.073 | 0.208 |
| NK_immature_NKG2A+ | 0.074 | 0.189 |
| MDSC3 | 0.078 | 0.182 |
| NK_mature_NKG2A+ | 0.081 | 0.138 |
| NKT_CD16+ | 0.081 | 0.153 |
| CD8+naive_CD57+ | 0.081 | 0.118 |

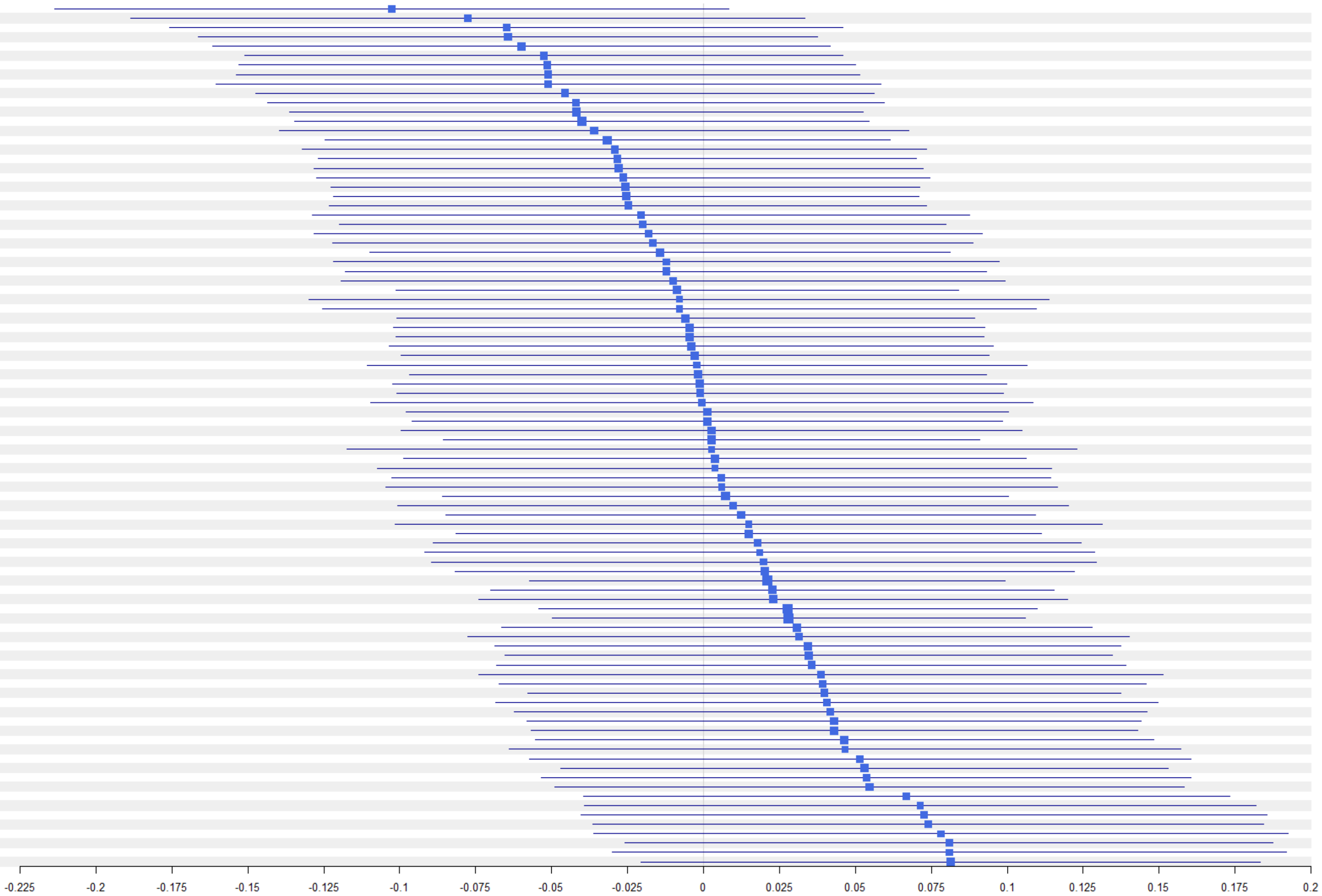

### 3D. AD, older age group

| Immune cell type | Beta | P value |
|---|---|---|
| CD8+LEM | -0.133 | 0.038 |
| NKT_CD3+ | -0.123 | 0.080 |
| CD8+LEM_CD57+ | -0.120 | 0.061 |
| CD8+CM_EM | -0.097 | 0.127 |
| NKT_CD56+ | -0.096 | 0.170 |
| CD3bright | -0.074 | 0.288 |
| Tcells_NKG2C+ | -0.067 | 0.340 |
| CD8+CD28+ | -0.066 | 0.297 |
| CD4+ME | -0.066 | 0.309 |
| CD8+CD27+ | -0.059 | 0.352 |
| CD4+ED_CD27-CD28+ | -0.059 | 0.357 |
| CD8+M_CD28+ | -0.048 | 0.451 |
| CD4+CD27+ | -0.047 | 0.458 |
| CD4+CD28+ | -0.043 | 0.500 |
| Bcells_NKG2C+ | -0.043 | 0.539 |
| M_I | -0.035 | 0.648 |
| CD4+TEMRA | -0.032 | 0.613 |
| M_C | -0.030 | 0.690 |
| CD8+ME | -0.030 | 0.631 |
| CD4/CD8_Ratio | -0.028 | 0.664 |
| CD8+TEMRA | -0.026 | 0.690 |
| Monocytes | -0.020 | 0.771 |
| Monocytes_CD14+ | -0.019 | 0.789 |
| CD4+SM | -0.018 | 0.782 |
| MDSC_CD14+CD15+ | -0.018 | 0.798 |
| CD8+ME_CD95+ | -0.016 | 0.800 |
| CD4+CD57+ | -0.015 | 0.808 |
| CD4+M_CD28+ | -0.015 | 0.815 |
| Tcells | -0.015 | 0.818 |
| CD8+M_CD27+ | -0.011 | 0.869 |
| NK_mature_NKG2C+ | -0.009 | 0.898 |
| CD8+CD57+ | -0.008 | 0.894 |
| CD14+CD15- | -0.008 | 0.906 |
| CD4+CM_EM | -0.008 | 0.897 |
| CD8+ED_CD27-CD28- | -0.007 | 0.909 |
| CD4+LEM | -0.007 | 0.912 |
| CD4+LD_CD28+ | -0.006 | 0.919 |
| Bcells | -0.006 | 0.926 |
| mDC | -0.005 | 0.945 |
| CD8+ED_CD27-CD28+ | -0.005 | 0.942 |
| Lin-CD14+HLA-DR+ | -0.003 | 0.969 |
| CD4+LEM_CD57+ | 0.000 | 0.999 |
| CD15high | 0.004 | 0.958 |
| CD4+ | 0.006 | 0.925 |
| CD4+ED_CD27-CD28- | 0.007 | 0.906 |
| MDSC4 | 0.012 | 0.864 |
| M_N | 0.015 | 0.860 |
| CD4+ME_CD95+ | 0.016 | 0.809 |
| MDSC2 | 0.017 | 0.807 |
| NK | 0.018 | 0.803 |
| CD8+LD_CD28+ | 0.018 | 0.780 |
| CD4+LD_CD27+CD28+ | 0.020 | 0.757 |
| MDSC2_w/oCD11b | 0.022 | 0.750 |
| sDC | 0.024 | 0.753 |
| CD3-CD16+ | 0.026 | 0.708 |
| CD4+ED | 0.027 | 0.672 |
| NKT_CD16+ | 0.029 | 0.683 |
| CD4+naive | 0.030 | 0.629 |
| CD4+LD | 0.034 | 0.595 |
| CD4+LD_CD27+ | 0.034 | 0.596 |
| CD8+PD-1+ | 0.034 | 0.585 |
| NK_immature_NKG2C+ | 0.038 | 0.596 |
| CD8+ | 0.038 | 0.551 |
| Tcells_NKG2A+ | 0.038 | 0.587 |
| CD4+M_CD27+ | 0.043 | 0.504 |
| Tcells_leuko | 0.044 | 0.524 |
| Lymphocytes | 0.046 | 0.506 |
| CD4+ED_CD27+CD28- | 0.046 | 0.463 |
| Lin-HLA-DR- | 0.047 | 0.505 |
| MDSC3 | 0.051 | 0.490 |
| CD8+ED_CD27+CD28- | 0.057 | 0.376 |
| Lymphocytes_living | 0.059 | 0.397 |
| CD8+ED | 0.059 | 0.331 |
| DC | 0.060 | 0.421 |
| CD8+naive | 0.063 | 0.307 |
| CD8+LD_CD27+ | 0.069 | 0.288 |
| Lin-HLA-DR-cross | 0.070 | 0.330 |
| CD8+LD_CD27+CD28+ | 0.073 | 0.242 |
| CD14-CD15- | 0.074 | 0.285 |
| CD3-CD16- | 0.074 | 0.292 |
| CD8+ME_CD57+ | 0.077 | 0.231 |
| CD8+SM | 0.077 | 0.230 |
| NK_immature | 0.079 | 0.265 |
| CD8+LD | 0.080 | 0.216 |
| NK_immature_NKG2A+ | 0.086 | 0.223 |
| MDSC1 | 0.087 | 0.221 |
| NK_mature | 0.095 | 0.181 |
| CD4+ME_CD57+ | 0.100 | 0.118 |
| CD4+naive_CD57+ | 0.102 | 0.112 |
| Bcells_NKG2A+ | 0.108 | 0.139 |
| CD8+naive_CD57+ | 0.156 | 0.015 |
| NK_mature_NKG2A+ | 0.158 | 0.024 |

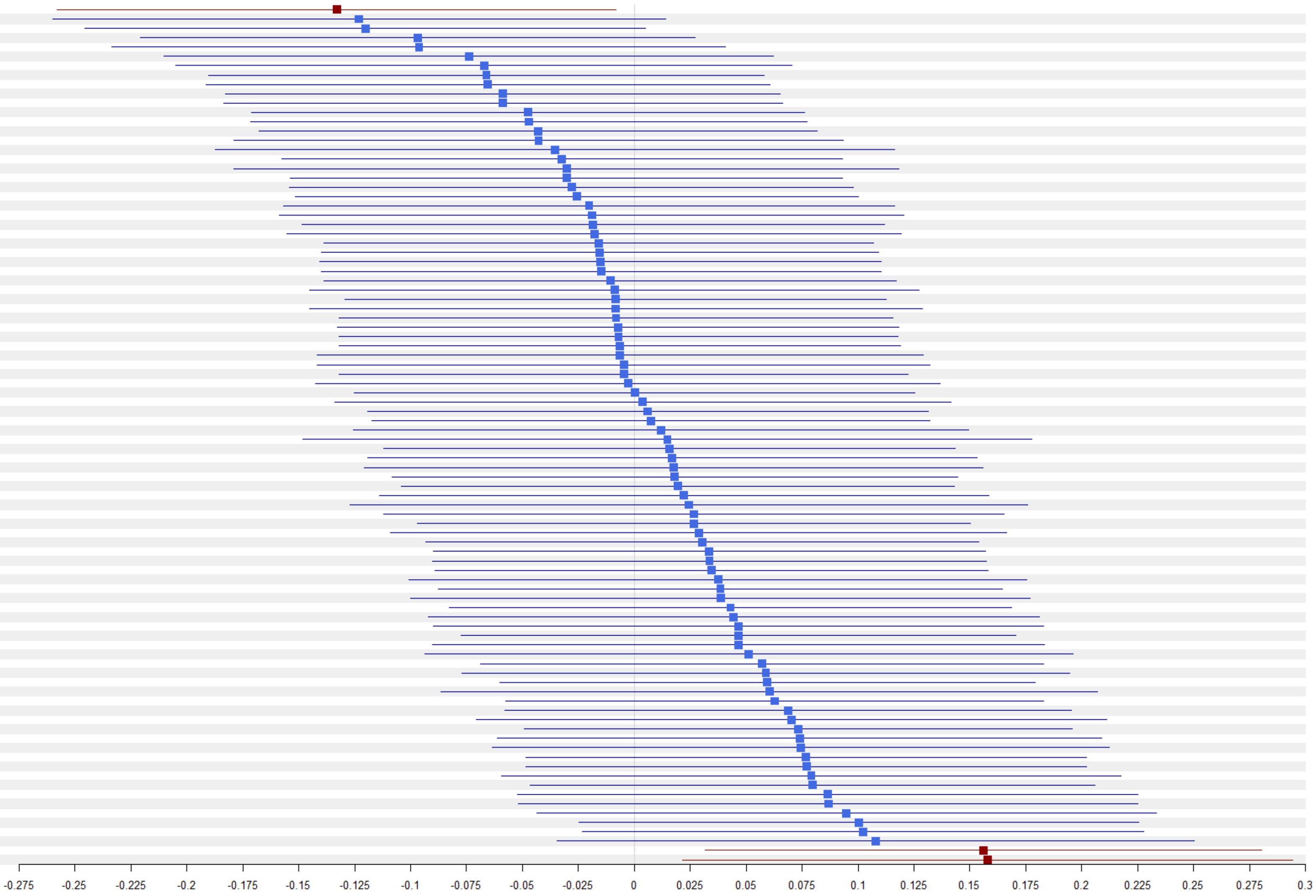
